# Emergence of SARS-CoV-2 variants of concern in the pediatric population of the United States

**DOI:** 10.1101/2021.05.22.21257660

**Authors:** Jennifer Dien Bard, Moiz Bootwalla, Amy Leber, Paul Planet, Ahmed M. Moustafa, Rebecca Harris, Pei Ying Chen, Lishuang Shen, Dejerianne Ostrow, Dennis Maglinte, Jessica Flores, Roy Somak, Sarangarajan Ranganathan, Elizabeth Perlman, Xiaotian Zheng, Rangaraj Selvarangan, Dithi Banerjee, Meghan Delaney, Joseph Campos, Drew G. Michael, Eric Vilain, Jonathan LoTempio, James Dunn, Sarah Jung, Samuel R. Dominguez, Alexander R. Judkins, Xiaowu Gai

## Abstract

The evolution of SARS-CoV2 virus has led to the emergence of variants of concern (VOC). Children, particularly <12 years old not yet eligible for vaccines, continue to be important reservoirs of SARS-CoV-2 yet VOC prevalence data in this population is lacking. We report data from a genomic surveillance program that includes 9 U.S. children’s hospitals. Analysis of SARS-CoV-2 genomes from 2119 patients <19 years old between 03/20 to 04/21 identified 252 VOCs and 560 VOC signature mutations, most from 10/20 onwards. 75.00% of the VOC signature mutations detected were in children <12 years old, including 32 cases of B.1.1.7 and 346 of B.1.419/B.1417. From 02/21 to 04/21, B.1.1.7 prevalence increased from 1.28% to 72.22% corresponding with the decline of B.1419/B.1417 from 52.57% to 5.56% at one institution. There continues to be a need for ongoing genomic surveillance, particularly among young children who will be the last groups to be vaccinated.

## Main text

The COVID-19 pandemic has provided a unique opportunity to observe the emergence and evolution of variants of concern (VOC) and variants of interest (VOI) through genomic surveillance. These VOCs were found to have significantly increased transmissibility and potential for immune escape(1,2). Continued high transmission rates in the U.S. increases concern for domestic VOCs such as the recently reported B.1.1.7 sub-lineage with spike protein D178H and membrane protein V70L mutations(3). Rates of vaccination are also slowing in the U.S. with 20-30% of people declining. In addition, children <12 years of age currently remain ineligible for vaccination. Further complicating this picture is prolonged COVID-19 infection in immunocompromised children with increased mutation rates, serving as potential reservoir of novel VOCs(4). Therefore, early detection of VOCs/VOIs and mutations in children is critical for ending the pandemic. However, to date pediatric populations are not the focus of genomic surveillance efforts.

We established a pediatric SARS-CoV-2 genomic surveillance program including 9 geographically diverse children’s hospitals in the U.S (**Supplementary Figure 1**) that serve a socioeconomically diverse and underrepresented population. This cohort is also temporally diverse, comprising samples collected from over a 13-month timeframe (03/20 to 04/21) affording the opportunity to observe the emergence of mutations over time. Whole genome sequencing (WGS) as previously described(5) was performed on 2119 SARS-CoV-2 positive samples detected from patients less than 19 years old. This consisted of all SARS-CoV2 positives from 1 pediatric institution (Children’s Hospital Los Angeles [CHLA], N=1669) and convenience samples from the remaining 8 institutions (N=450). The median age of patients was 6.9 years (5 days–18 years). Reasons for SARS-CoV-2 testing include symptomatic presentation, pre-procedural asymptomatic screening, and surveillance.

We analyzed the viral genomes to identify key mutations associated with known VOC/VOI (N501Y, E484K, L452R)(6,7,8) and identified 560 key mutations and 525 VOCs total (**Supplementary Table 1)**. This analysis demonstrated a geographically specific pattern for emergence of VOC/VOI in children best illustrated by the B.1.429/B.1.427 or so-called California variant. Of the 887 isolates tested between 03/20 to 10/20, only 11 (1.24%) isolates harboring one of the three mutations were identified. Specifically, at CHLA, the B.1.429/B.1.427 lineages (L452R mutation) were first detected on 09/20 (N=6, 9.52%), increasing with the Fall surge on 11/20 (N=26, 14.53%), peaked on 01/21 (N=215, 52.57%), followed by a significant decline on 04/21 (N=1, 5.56%). For the period of 11/20 to 04/21, B.1.429/B.1.427 lineages accounted for 443 patients, or 98.01% at CHLA. This is consistent with data derived primarily from adults which report that B.1.429/B.1.427 lineage is 20% more transmissible and demonstrates increased resistance to certain monoclonal antibodies(6). In contrast, of 199 isolates available for the same period from 4 other institutions, only 7 (3.52%) were identified as B.1.429/B.1.427 lineages (**Supplemental Table 2**).

Beginning on 11/20 we also observed a significant increase in N501Y mutation in the spike protein, a key mutation found in the VOCs B.1.1.7, B.1.351 and P.1 lineages. At CHLA, the emergence and rapid increase of B.1.1.7 from 1.28% (1/78) on 02/21 to 72.22% (13/18) on 04/21 corresponded with the decline of B.1.429/B.1.427 over the same period (**Figure 1, supplemental Table 1**). Thirty-one additional B.1.1.7 isolates from 3 other institutions (CHOP, LCH and NCH) were observed during the same period highlighting that children are reservoirs of this VOC and that B.1.1.7 effectively outcompeted B.1.429/B.1.427 in a region where the latter had previously been well established.

**Figure 1.**
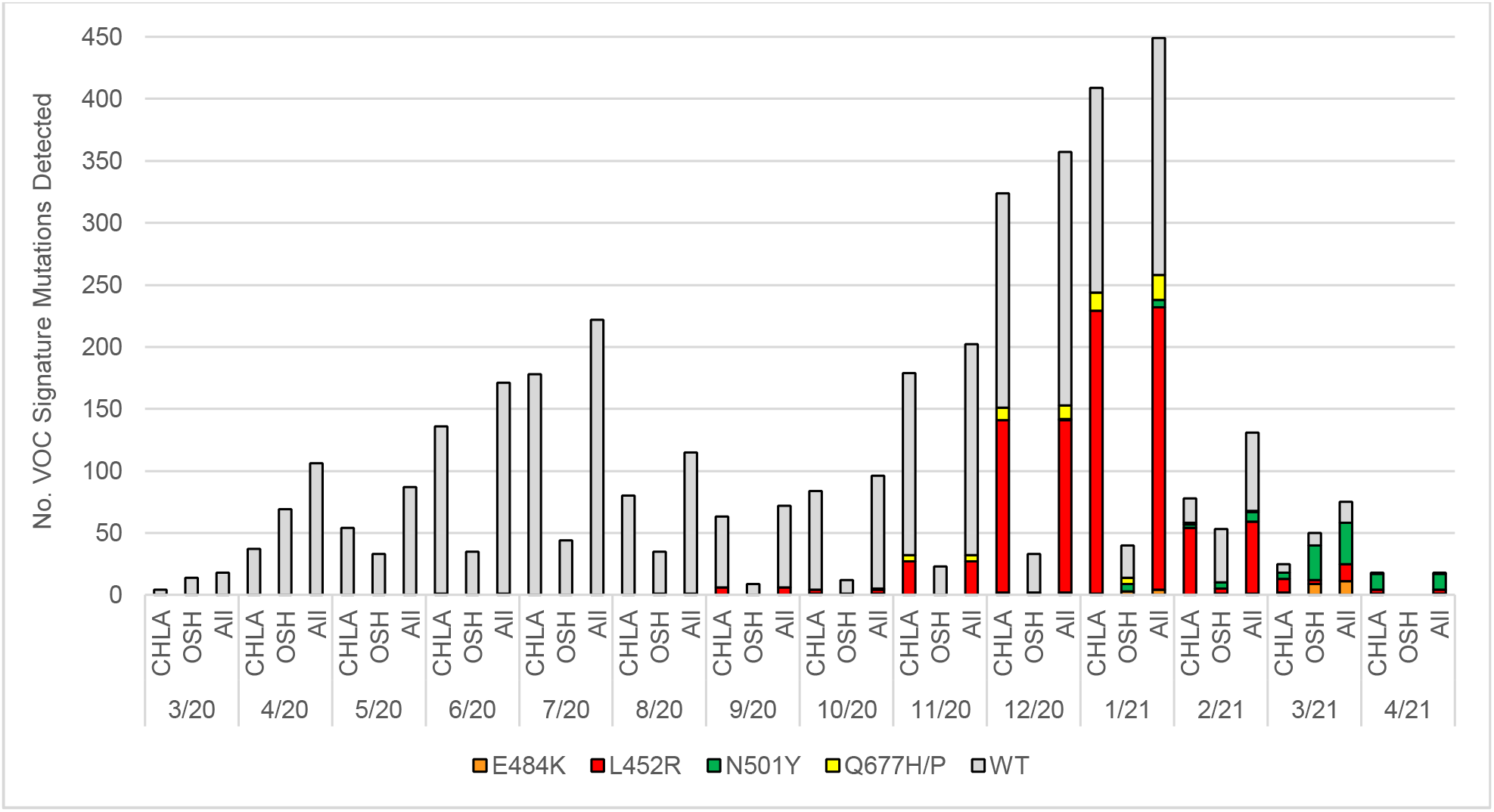
Key mutations in SARS-CoV-2 isolates tested over 13-month period. Prevalence of N501Y, E484K, L452R and Q677H/P identified by SARS-CoV-2 whole genome sequencing (WGS) compared to wild-type (WT) SARS-CoV-2 isolates were tested by WGS in pediatric patients <19 years of age seen at 9 Children’s Hospital in the U.S. Wild-type isolates were defined as a SARS-CoV-2 isolate that does not carry any VOC signature mutations. All samples (ALL) were compared to samples tested at Children’s Hospital Los Angeles (CHLA) and samples tested at the 8 other institutions (OSH).

Likewise, another mutation in the spike protein, E484K, which has been reported to allow for escape from neutralizing antibodies and found in multiple VOCs/VOIs including P.1, P.2, B.1.351, and B.1.526 lineages, was detected in 21 pediatric patient samples, including P.1 (N=2), P.2 (N=3), B.1.351 (N=1) and B.1.526 (N=8) (**Supplemental Table 2**). Interestingly, we identified one B.1.1.7 isolate that also harbored the E484K spike mutation(9). Finally, over the period 10/20 to 04/21 we identified several isolates harboring the Q677H or Q677P mutation with only a modest increase (1.04 to 5.56%). These mutations were reported from Louisiana and New Mexico but appears to be more widespread already in other states(10).

A key finding is that nearly three-quarters (75.00%, 420/560) of the key mutations detected were recovered from children <12 years of age. This includes B.1.1.7 (N=32), P.1 (N=2), B.1.351 (N=1), and B.1.429/B.1.427 (N=346) (**Supplemental Table 2**). At a time of uncertainty about the value of continued non-pharmacological interventions, this highlights the very real risk posed by VOCs/VOIs to the yet unvaccinated/low rates of vaccination in the pediatric population. Our results provide clear evidence of the emergence of VOCs/VOIs in pediatric patients across diverse geographies and socioeconomic populations in the US. In conjunction with growing evidence for the risk of emergence of VOCs among chronically infected immunocompromised children, this report highlights the need for ongoing genomic epidemiological surveillance among pediatric populations who will be among the last groups to receive vaccination and who are key to ending this pandemic.

## Data Availability

All genomic data were uploaded to GISAID. The corresponding accessions are available upon request from the corresponding author.

## Acknowledgement

The work was partially funded by The Saban Research Institute at Children’s Hospital Los Angeles intramural support for COVID-19 Directed Research (X.G. and J.D.B.). We would like to acknowledge the staff members of the Clinical Microbiology and Virology laboratory and Center for Personalized Medicine at Children’s Hospital Los Angeles for conducting SARS-CoV-2 RT-PCR and WGS.

## Figure Legends

**Supplementary Figure 1.**
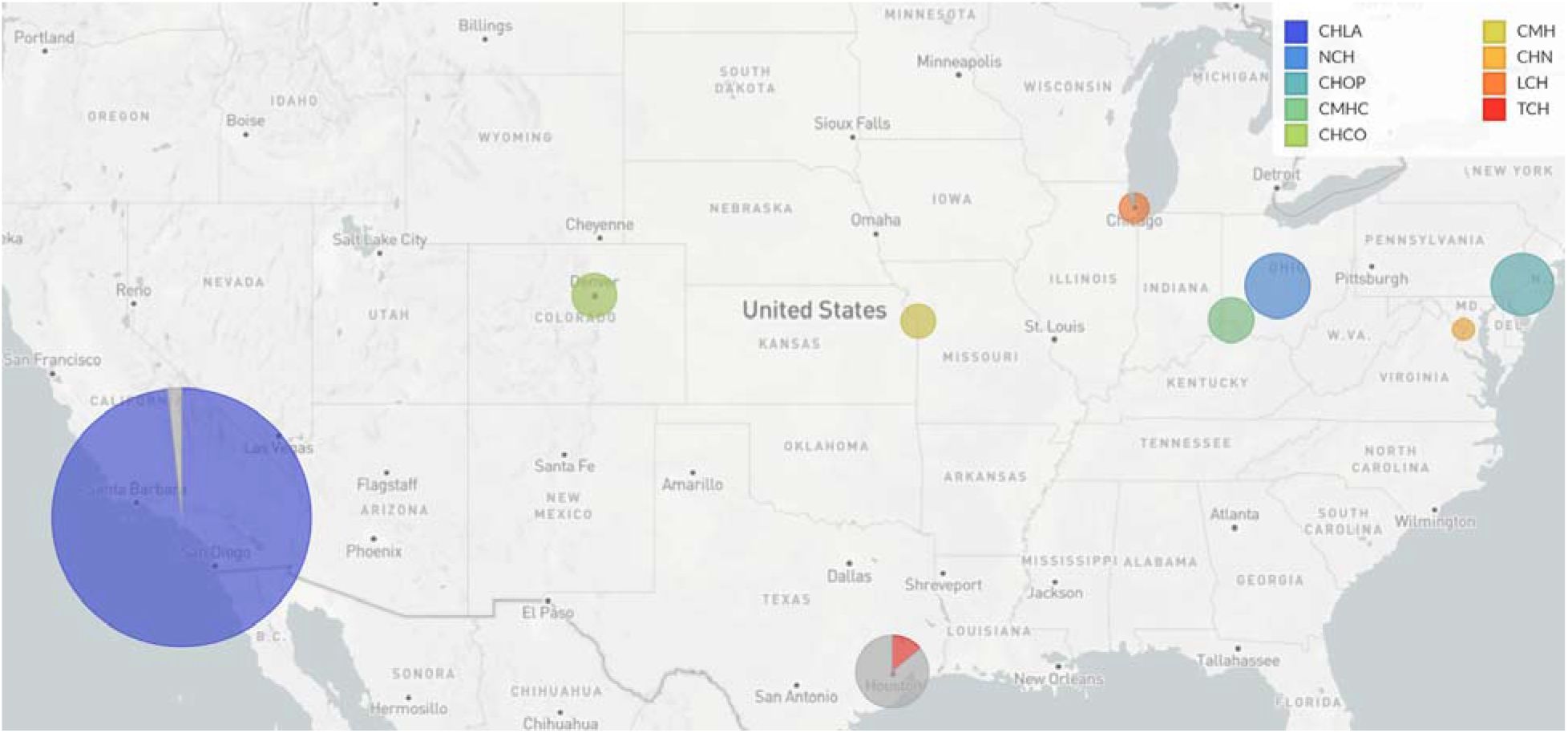
Geographical distribution of 9 Children’s Hospitals in the U.S. Whole genome sequence and genomic analysis was performed on a total of 2119 SARS-CoV-2 isolates from 9 geographically diverse pediatric institutions: 1. Children’s Hospital Los Angeles; 2. Children’s Hospital Colorado; 3. Texas Children’s Hospital; 4. Children’s Mercy Kansas City; 5. Lurie Children’s Hospital; 6. Cincinnati Children’s Hospital; 7. Nationwide Children’s Hospital; 8. Children’s National Hospital; 9. Children’s Hospital of Philadelphia. The pediatric institutions are highlighted (in color) were merged with subsampled genomes carrying identical haplotypes from the same cities (in grey). The dot sizes are representative of the resulting number of genomes per city.

**Supplementary Table 1**. Prevalence of Variant of Concern Signature Mutations Identified from 2119 Pediatric Patients 18 years and Younger from Nine Children’s Hospital

**Supplementary Table 2**. Prevalence of Variant of Concern Detected in Pediatric Patients 18 years and Younger from Five Children’s Hospital

